# A roadmap for implanting microelectrode arrays to evoke tactile sensations through intracortical microstimulation

**DOI:** 10.1101/2024.04.26.24306239

**Authors:** John E Downey, Hunter R Schone, Stephen T Foldes, Charles Greenspon, Fang Liu, Ceci Verbaarschot, Daniel Biro, David Satzer, Chan Hong Moon, Brian A Coffman, Vahab Youssofzadeh, Daryl Fields, Taylor G Hobbs, Elizaveta Okorokova, Elizabeth C Tyler-Kabara, Peter C Warnke, Jorge Gonzalez-Martinez, Nicholas G Hatsopoulos, Sliman J Bensmaia, Michael L Boninger, Robert A Gaunt, Jennifer L Collinger

## Abstract

Intracortical microstimulation (ICMS) is a method for restoring sensation to people with paralysis as part of a bidirectional brain-computer interface to restore upper limb function. Evoking tactile sensations of the hand through ICMS requires precise targeting of implanted electrodes. Here we describe the presurgical imaging procedures used to generate functional maps of the hand area of the somatosensory cortex and subsequent planning that guided the implantation of intracortical microelectrode arrays. In five participants with cervical spinal cord injury, across two study locations, this procedure successfully enabled ICMS-evoked sensations localized to at least the first four digits of the hand. The imaging and planning procedures developed through this clinical trial provide a roadmap for other brain-computer interface studies to ensure successful placement of stimulation electrodes.

## Introduction

Individuals with tetraplegia consistently report restoration of hand and arm function as a top rehabilitative priority^1,2^. To address this need, researchers are developing brain-computer interfaces (BCIs) that can bypass the injured spinal cord to enable control of a robotic arm^3–9^ or to restore movement via functional electrical stimulation (FES)^6,10^. However, motor control alone is not sufficient to fully restore arm and hand function or the ability to interact with the world. Somatosensory feedback is essential to maintained motor function^11^. Indeed, the loss of somatosensation drastically impairs hand and arm function even when motor pathways remain intact^12^. Tactile feedback enables manipulation and exploration of objects and provides affective qualities that are unavailable in a purely motor BCI^13^. Therefore, we aim to provide intuitive tactile sensory feedback through intracortical microstimulation (ICMS) in somatosensory cortex with microelectrode arrays in the post-central gyrus.

The intuitiveness of the sensory feedback is essential for maximizing both the embodiment of the BCI as well as its functionality. ICMS of somatosensory cortex has arisen as a promising approach to evoke intuitive sensory feedback as part of a BCI^14–17^, as the location and intensity of the sensations can be systematically controlled. Somatosensory cortex is also an ideal candidate for surgical targeting as it is organized somatotopically, with discrete regions receiving the sensory inputs from specific parts of the body^18–20^. In particular, the hand representation progresses from the thumb, laterally in somatosensory cortex, through the digits to the little finger medially^21^. The surface of the post-central gyrus, the section of somatosensory cortex accessible to planar microelectrode arrays, mostly consists of Brodmann’s area 1^22^. In area 1, fingertips are represented at the border with area 2 – the posterior edge of area 1 – and finger bases are represented at the border with area 3b – the anterior edge of area 1^23^. The latero-medial organization from the thumb to the little finger in somatosensory cortex was originally observed by Penfield using intraoperative electrical stimulation of humans^24,25^ and has been confirmed in multiple species^26–30^ including non-human primates^18–20,31^. This somatotopic finger organization can also be observed in humans using non-invasive imaging techniques – functional magnetic resonance imaging (fMRI) and magnetoencephalography (MEG)^32–40^. Importantly, several studies have demonstrated highly preserved finger representations in people who were deafferented through amputation^41–44^ or spinal-cord injury (SCI)^45^; the two groups of people most likely to benefit from BCIs to restore upper limb function.

As part of a multisite clinical trial, we implanted ten intracortical microelectrode arrays (Blackrock Microsystems, Inc., Salt Lake City, UT) in the post-central gyrus of five study participants (2 arrays in each) with chronic SCI to provide ICMS-based tactile sensory feedback. Here we present a roadmap to generate and execute a successful plan for evoking sensations localized to the desired digits. We developed this rigorous plan after a misplaced implant that resulted in no sensations during stimulation^46^. We used fMRI and/or MEG^47^ to reliably identify the regions of somatosensory cortex that represent touch sensations for digits despite differences in the residual sensory function of the participants. Presurgical data analysis and planning of array locations were performed by a broad study team with expertise in neurosurgery, neuroscience, engineering, and spinal cord injury medicine. Using this plan and a structured surgical workflow that required input from multiple team members, we successfully identified implant locations such that ICMS evoked sensations in the digits in all five participants. We expect that this workflow can define a pathway to broader use of BCIs that provide intuitive, somatotopically-matched sensory feedback.

## Results

### Participants

All five participants (C1, C2, P2, P3, and P4) presented with paralysis of the right hand and some amount of deafferentation due to cervical level SCI (and brachial plexopathy in C2). Table 2 describes the clinical aspects of the participants. C1 retained deep sensation with diminished light touch across his whole right hand. P2 and P3 were insensate across the ulnar region (middle to little fingers) of the right hand but retained diminished light touch and deep sensation on the radial side. C2 and P4 were insensate across the entirety of their right hands.

### Functional Imaging

To localize the hand representation of each participant, we used a combination of fMRI and MEG. Four participants completed the fMRI task in which they were instructed to attempt to perform a sequential finger tapping task - the travelling wave paradigm^36,41^. Three participants (including 2 who completed the fMRI task as well) completed the MEG tasks in which they alternated attempted tapping of individual fingers and rest periods while viewing a representation of the desired movement. While the participants were unable to physically make the movement, we identified representations of at least three digits on the post-central gyrus, which was accessible to microelectrode arrays, in each (Figure 1). The hand representation of somatosensory cortex is often assumed to be adjacent to the hand knob – an identifiable landmark of the precentral gyrus (motor cortex) that is preferentially activated during hand movements and is generally anatomically distinct^48^. We found, however, that the position of the sensory representations relative to the hand knob varied by up to 25 mm along the mediolateral axis across participants depending on the digit (Table 1, Supplementary Figure 1). The location variability is more than 6x the size of the NeuroPort microelectrode arrays (2.4x4mm, Blackrock Neurotech, Salt Lake City, UT), thus rendering the hand knob an insufficient targeting landmark.

**Figure 1.**
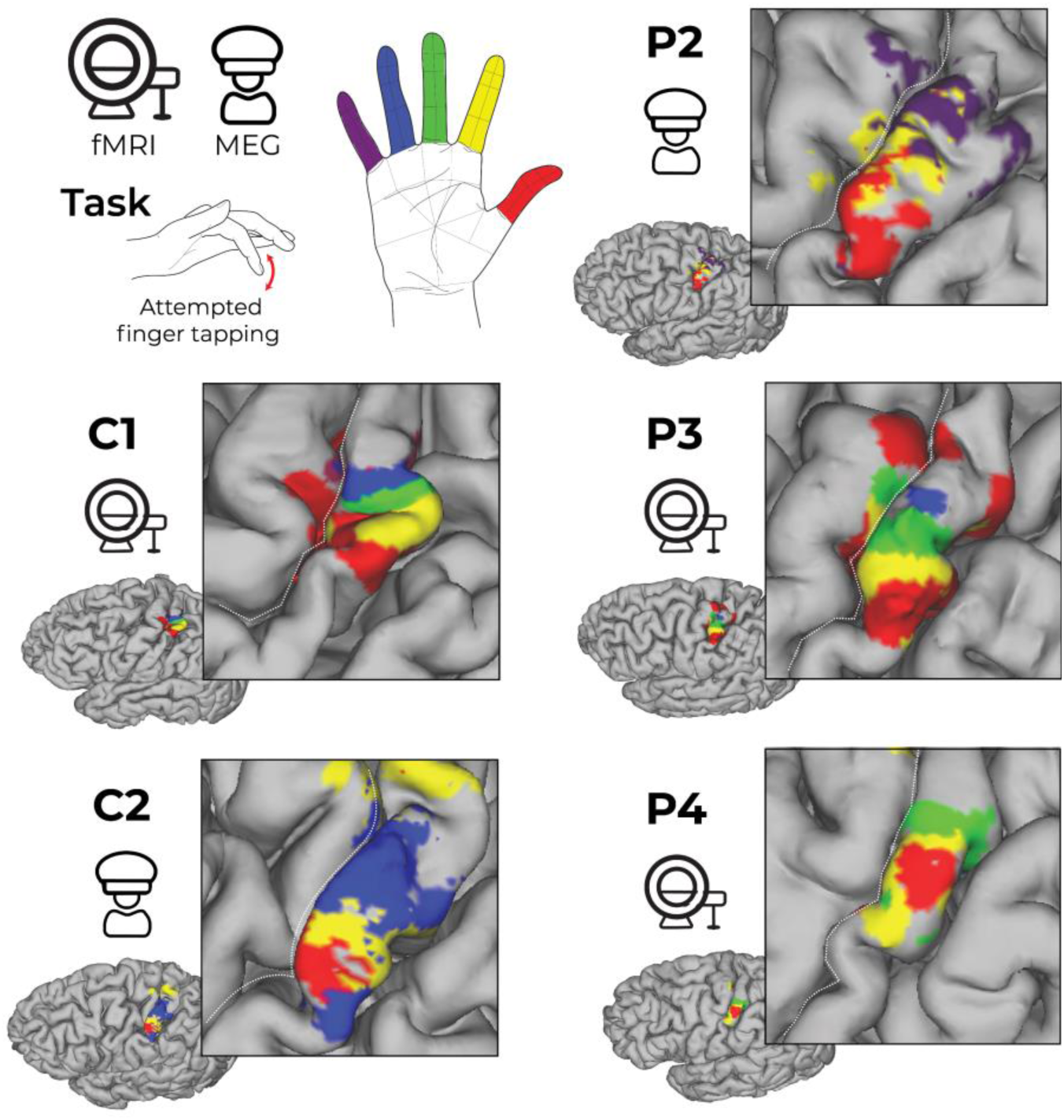
Functional activity projected on the post-central gyrus of each participant. Each map displays the significant activity in the somatosensory cortex from the mapping tasks during fMRI or MEG. For C1, P3, and P4, all digits were tested. For P3, no significant modulation was found for the little finger. For P4, no significant modulation was found for the ring or little fingers. For P2, only thumb, index, and pinky fingers were tested. For C2, only thumb, index, and ring fingers were tested. Finger visualization thresholding for fMRI was z-statistic > 3.1, while for MEG z-statistic > 50% of max was used, see online methods for details.

**Table 1.** Lateral distances of digit activity from the hand knob. The distance (mm) along the medial-lateral axis from the hand-knob to the peak activity for each digit that had significant activity measured on fMRI or MEG. Notice that individual digits could be separated by many mm in different participants. Absent values indicate digits without a significant functional map. Note that D3 and D5 were not tested for participant C2 and D4 and D4 were not tested for participant P2.

| Participant | Digit |  |  |  |  |
| --- | --- | --- | --- | --- | --- |
|  | D1 | D2 | D3 | D4 | D5 |
| C1 | 19.7 | 10.3 | 5.6 | 4.1 | 1.5 |
| C2 | 16.2 | 3.2 |  | 6.2 |  |
| P2 | 22.0 | 11.8 |  |  | 6.1 |
| P3 | 15.3 | 12.3 | 10.9 | 6.2 |  |
| P4 | 19.9 | 28.1 | 15.7 |  |  |

**Table 2.** Participant demographics. SCI = spinal cord injury; ASIA=American Spinal Injury Association; fMRI=functional magnetic resonance imaging; MEG=magnetoencephalography

|  | C1 | C2 | P2 | P3 | P4 |
| --- | --- | --- | --- | --- | --- |
| **Age** | 50s | 60s | 20s | 20s | 30s |
| **Gender** | Male | Male | Male | Male | Male |
| **Injury** | SCI (C4 ASIA D) | SCI (C4 ASIA D) and right brachial plexopathy | SCI (C5 ASIA B) | SCI (C6 ASIA B) | SCI (C4 ASIA A) |
| **Years since injury** | 35 | 4 | 10 | 12 | 11 |
| **Filament testing** | Deep sensation, but diminished light touch in the right hand | Insensate across whole right hand | Insensate in the ulnar hand region, retained diminished light | Insensate in the ulnar hand region, retained diminished light | Insensate across whole |
|  |  |  | touch and deep<br>sensation on<br>the radial side | touch and deep<br>sensation on<br>the radial side | right<br>hand |
| **Scans**<br>**Performed** | fMRI | fMRI and<br>MEG | MEG | fMRI | fMRI and<br>MEG |

We found that fMRI was not feasible in some cases. P2 reported a warming sensation during a previous fMRI scan (unrelated to the current study) that may be related to his implanted spinal fixation hardware, so an fMRI study was not completed. Two other participants (C2 and P4) experienced claustrophobia leading to excessive head movement during the fMRI task resulting in noisy data. Therefore, an MEG scan was completed for P2, P4, and C2. During the MEG, the number of fingers tested was limited due to time constraints. Despite the fundamental differences between fMRI and MEG, the functional maps closely matched between the two imaging modalities in the two participants where both modalities were used, with the center of gravity for each digit varying by between 3.1 and 6.8 mm (Supplementary Figure 2). Ultimately, fMRI and MEG were able to map the location of finger representations across participants with SCI of varying levels, completeness, and time since injury.

### Surgical planning

Once functional imaging results were obtained, the study team (including both the University of Pittsburgh and University of Chicago clinical trial teams) planned the preferred array implant locations. The planning process was parallelized at multiple points (Figure 2) to avoid groupthink^49^ and ensure confidence in final decision making. For all participants – except P2, whose implant preceded the addition of a second clinical trial site – both study sites initially analyzed the functional imaging data separately to generate functional maps. Once both sites generated functional maps, any differences were identified and the team members responsible for generating the functional maps collaborated to determine where any errors were made and resolve them. Once both sites agreed on the functional map it was distributed to 7 or 8 senior researchers, including neuroscientists, neurosurgeons, and physiatrists, to make array placement plans independently. When creating individual placement plans, we considered which areas most strongly represented each finger, where the finger representations bordered or overlapped to allow one array access to both fingers, where the gyrus appeared flat enough for array placement, and where large blood vessels were located (based on a structural MRI with contrast, Figure 2b). After all independent plans were ready, these plans were compiled and only then were they discussed together (e.g., Supplementary Figure 3). The full group generated a consensus plan for array placement locations (Figure 2b, Figure 3a), including backup locations for array placement in case the initial site was found to be unsuitable for array placement during surgery. The planned locations were uploaded to a surgical navigation system (StealthStation, Medtronic, Minneapolis MN; Brainlab, Munich, Germany; or ROSA®, Zimmer Biomet, Warsaw, Poland) and coregistered with a structural T1-weighted MRI with contrast and 1 mm slice thickness, which identified large blood vessels (Figure 2b).

**Figure 2.**
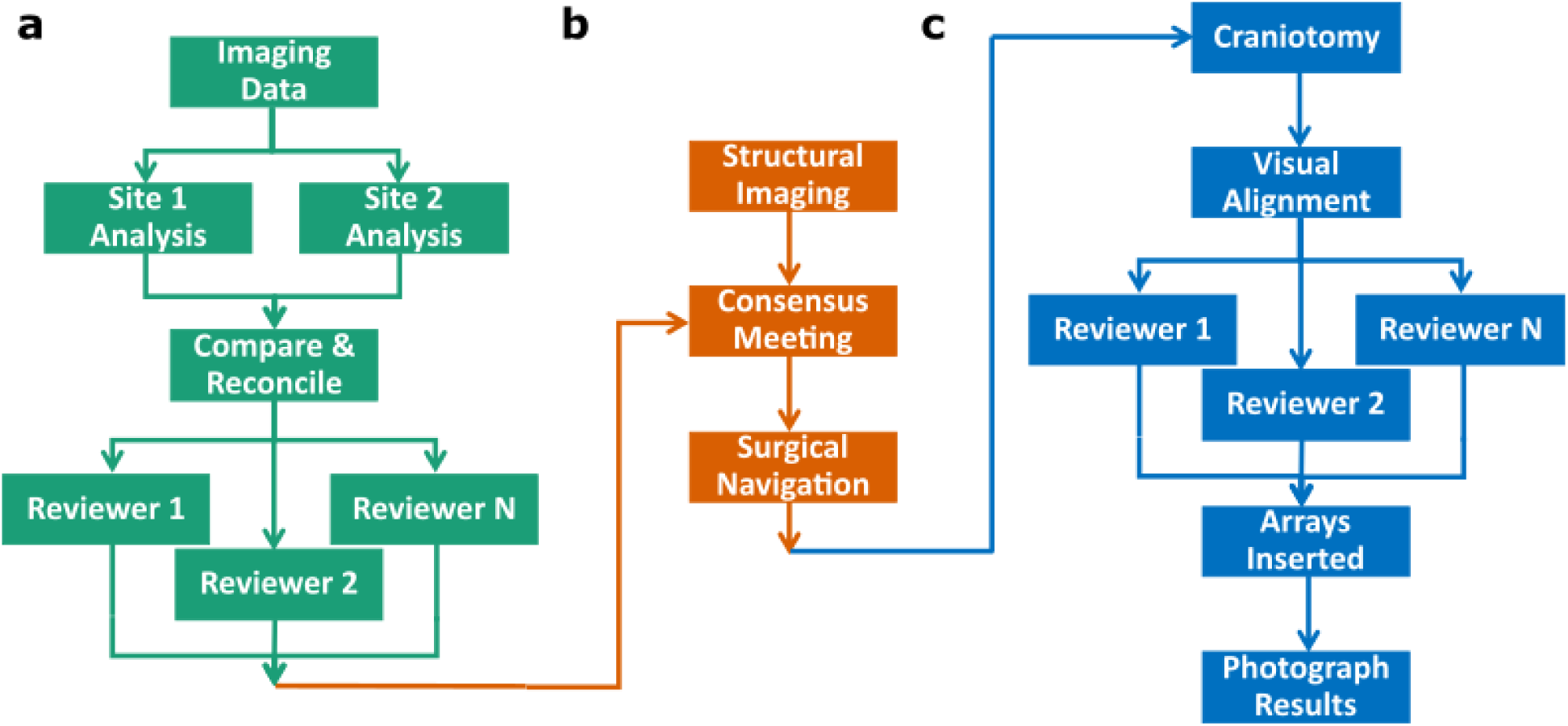
Preoperative and intraoperative array placement planning procedures. a) Functional imaging pipeline, including data collection, parallel analysis of raw data, and parallel selection of preferred implant locations. b) Preoperative preparations, including a structural MRI with contrast, a meeting of team members to develop a consensus implant plan, and the translation of that plan to surgical navigation software. c) Intraoperative procedures, using the plan to expose the correct brain area, visually confirming the anatomical location, and determining the final implant locations after accounting for any new information. Post insertion confirmation of location is done using photographs taken during surgery.

**Figure 3.**
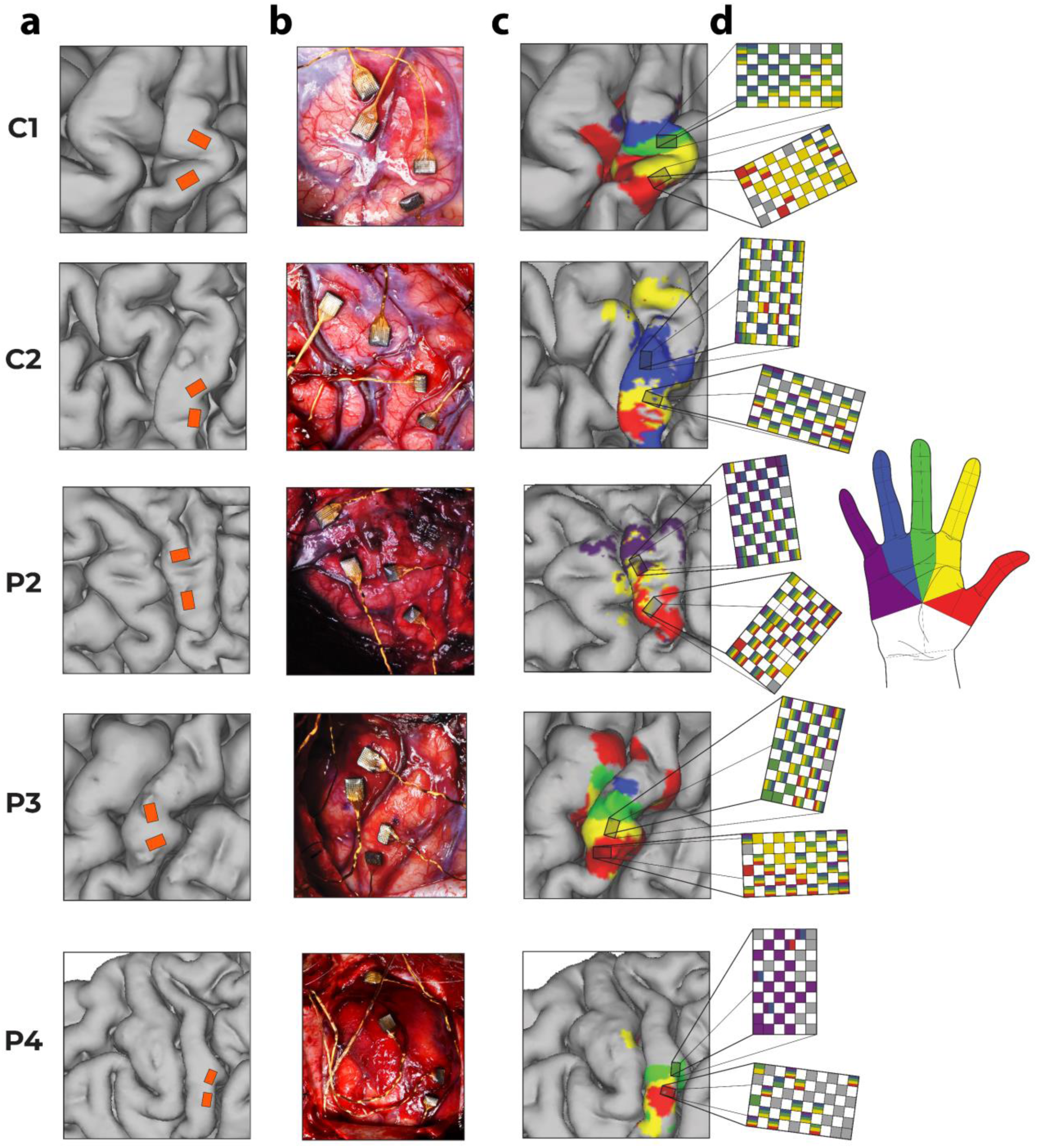
Locations of the planned and inserted electrode arrays with the functional maps and their corresponding projected fields. a) Consensus presurgical plan for array locations. b) Post-implant intraoperative photo of electrode array locations. c) Shaded boxes indicate the final array locations on the pre-surgical functional maps described in Figure 1. d) The expanded views of the sensory arrays are colored based on the location of the projected fields for each electrode, as illustrated on the hand, which is where the participant experienced a sensation upon ICMS of that electrode.

### Surgical Approach

Using stereotactic image guidance in the operating room, the craniotomy location was planned based on the desired array locations within the navigation system. The neurosurgeon consulted with the other team members to confirm the opening would be the correct size and location to accommodate the planned array insertions and percutaneous pedestal locations, before performing the craniotomy. Once the craniotomy and dura resection were completed, a navigation wand or robotic arm was used to locate the implant site on the exposed cortex based on the neuroimaging data. Then we took photographs of the exposed cortex, and the researchers present in the operating room compared these photographs to the implant plan based upon the structural MRI (via digital overlay). The researchers visually aligned the photographs with the pre-surgical plan based on large landmarks, such as the central sulcus, major vessels, and gyrification patterns. This process was completed in parallel to create redundancy and ensure confidence when transferring the digital plan to the exposed cortex. A formal time-out was performed after the parallel reviews, during which all researchers came to a consensus regarding the mapping of exposed cortex onto the planned locations (Figure 2c). Despite planning based on structural and functional imaging, there are factors that influence array placement and are only evident during surgery. These factors include small blood vessels that are not visible in the structural MRI, excessive curvature of the cortex not appreciated in the structural MRI, and viable paths for wire-bundle placement. The team then reconsidered the original array placement plan based on the information available at the time of implant to determine the final location for array placement. Formal consensus regarding the planned array location was obtained prior to insertion. After array insertion, we again took photographs (Figure 2c, Figure 3b) to allow for post-surgical coregistration with planned implantation locations (Figure 3c). Final array locations were consistent from the consensus plan to the implanted result (Figure 3a and Figure 3c, respectively). The one exception was the medial array in C2, due to extensive vasculature on the posterior half of the post-central gyrus. Array orientations, however, frequently changed from the plan during surgery, due to vasculature or array wire pathway requirements.

### Projected Fields

After the participants recovered from the surgery and began attending testing sessions in the lab, we delivered ICMS to each electrode and surveyed the participants to determine the location of any evoked sensations – the “projected fields”. Projected fields were located on the hand for all electrodes (for which there was a percept) in all participants, spanning from the thumb to the little finger and from the metacarpophalangeal joint (MCP) to the fingertip on both the palmar and dorsal sides of the hand. ICMS only ever produced sensations on the hand. The details of projected field sizes and distributions in participants C1, P2, and P3 were reported in Greenspon et al. 2023^16^. Here, we compare the correspondence between the expected digit representation obtained with functional imaging and the location of sensation evoked from stimulation of those arrays (Figure 3). In all cases except for P4’s medial motor array we had 100% correspondence— meaning that when the functional map predicted a particular digit’s representation on an array, ICMS of at least 1 electrode on that array evoked a sensation on that digit. The medial array in P4 overlaid the functional maps for D2 and D3 but resulted in sensations on D4 and D5, which did not have significant fMRI activity anywhere on the gyrus. Some electrodes evoked sensations on digits other than those predicted by functional imaging, but this was less common than evoked sensations on the predicted digit (Supplementary Table 1).

Though the imaging protocol was not designed to separate representations of individual finger segments (i.e., the MCP, proximal phalanx, middle phalanx or distal phalanx), we observed that most projected fields on single electrodes were confined to a single segment or two adjacent segments, even when felt on multiple digits. The location of electrodes’ projected fields on the proximal-distal axis tended to cluster, e.g. most electrodes in C1 had distal projected fields, and the lateral array in P2 had one edge with distal projected fields while the rest of the electrodes had proximal projected fields (Figure 4). However, despite this clustering the was no clear relationship between an electrode’s anterior-posterior location and the location of its projected field on the proximal-distal axis of the hand across participants.

**Figure 4.**
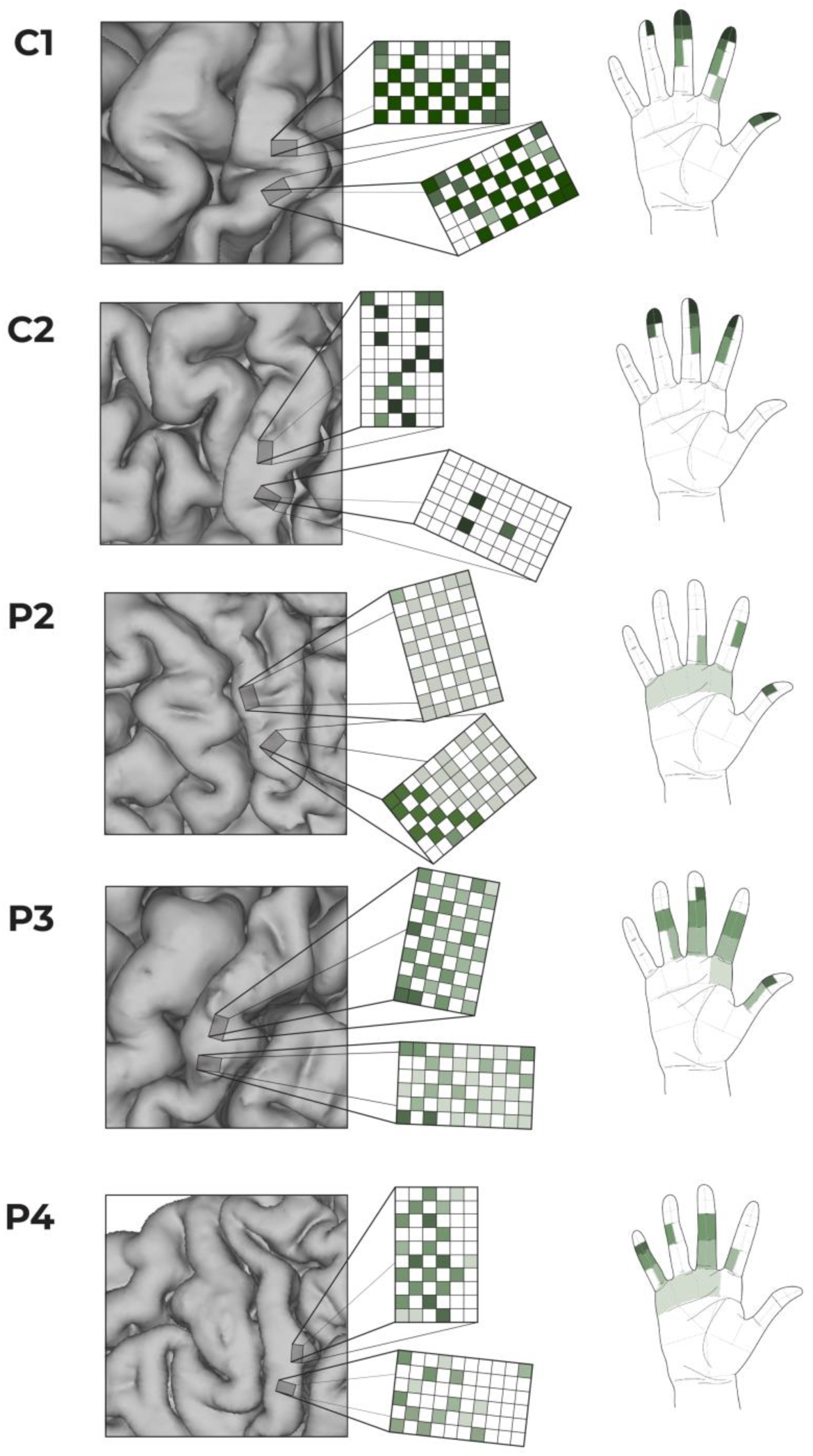
Comparison of electrode location and the central location of the projected field along the proximal-distal axis of the hand. Darker shades indicate more distal projected fields. No consistent pattern emerges to correlate electrode location with the proximal-distal location of the projected fields.

## Discussion

We developed a framework for reliably targeting the hand representation in somatosensory cortex such that ICMS can be used to deliver tactile feedback for BCIs. The precise targeting in somatosensory cortex is of particular importance as the somatotopy is more rigid than in motor cortex^50^ and the lack of sensory input makes functional mapping more difficult in the target population. Non-invasive neuroimaging (fMRI and MEG) was used to create individualized functional maps of somatosensation during attempted finger movements. Then, we used a clear strategy based on maximizing communication and preparation, while minimizing individual biases, to create a strong pre-operative plan that could be applied in the operating room. We demonstrated that this approach consistently resulted in evoked sensations that map to the desired digits. This level of successful targeting and planning enabled closed-loop BCI systems that provided tactile feedback during grasping^15,17^.

### Generating Functional Maps

We generated functional maps of at least three digits with either fMRI or MEG for each of the five participants. The imaging data were collected while the subjects attempted to move individual fingers, even though their ability to generate those movements was limited and therefore they did not feel appropriate sensory feedback. Our results show substantial variability in the mediolateral location of the finger representation relative to the anatomical hand knob in motor cortex between participants, consistent with prior intraoperative mapping work that showed a standard deviation exceeding 5 mm for some digits^32^. A longitudinal fMRI study similarly showed consistent digit localization across time within subjects but variable localization across subjects^37^. Variability in the location of the sensory digit maps relative to the anatomical hand knob, highlights the importance of functional mapping in determining electrode placement. Further, anatomical variability^51^ can lead to challenges in consistently identifying the hand knob of motor cortex. Of note, while we did not have imaging data prior to their injuries, we saw that the expected somatotopic organization is well preserved up to 35 years post-injury, joining growing evidence that substantial reorganization of the somatosensory cortex does not occur after injury in adults^43,52–54^.

All participants tolerated an MRI with contrast for structural imaging; however, it was more challenging to obtain quality fMRI data due to claustrophobia, movement, or the potential risk of hardware heating when using 3T fMRI scan parameters. Digit mapping with fMRI requires a significant amount of time in the scanner (30-45 minutes) and requires active task participation. While fMRI is more widely available, MEG provided a viable alternative functional imaging method. The fact that both our fMRI and MEG functional maps aligned in two participants and that the MEG-only implant (P2) had well predicted projected fields demonstrated that both approaches are suitable for generating functional maps. While 90% of arrays evoked sensations on the expected digits, most arrays also evoked sensations on digits that were not mapped under them (Supplementary Table 1). This may be due to overly conservative thresholding of the functional maps, difficulty in evoking activity for certain digits during imaging, or stimulation of transiting axons versus local cell bodies. We recognize that functional neuroimaging may not be broadly available at medical centers, which could create challenges for translation of bidirectional BCI technology. However, as more data are collected we will gain knowledge of the typical location and size of sensory digit representations in people with tetraplegia. This could also inform the development of electrode technology that can broadly cover the entire hand representation and reduce the need for such precise electrode placement.

### Surgical Considerations

Other studies of ICMS in the somatosensory cortex have placed electrodes using intraoperative mapping^55,56^. We chose to use pre-operative functional mapping because the heterogeneity of deafferentation can make it difficult to interpret recorded cortical responses to peripheral stimulation; the main method of intraoperative mapping^55^. There is also an increased risk of autonomic dysreflexia among this patient population if they are awakened during the surgery^57,58^, preventing participants from verbally reporting the location of sensations during intraoperative stimulation of cortex^32,56^. Given these concerns, and risks associated with further prolonging the surgery, we believe functional imaging was a more versatile and safe way to determine where to place electrodes.

Successful targeting of the arrays is of particular importance due to the costs and risks involved with implantation. Indeed, our group’s first experience attempting to place stimulating arrays in somatosensory cortex was unsuccessful^46^, leading us to create this thorough and robust plan (Figure 2) to ensure accurate placement in the future. In that original implant, we did not have the ability to import functional neuroimaging maps into a surgical navigation system and instead relied on introperative anatomical navigation only. Given the variety of priorities during an implant, we had multiple team members with diverse backgrounds (neuroscientists, engineers, rehabilitative medicine clinicians and neurosurgeons) generate parallel plans. Each team member brings unique insights and priorities, resulting in different opinions on the precise placement and orientation before these parallel plans were consolidated (Supplementary Figure 3). In our experience, a unanimous consensus was always reached. However, it is important that there be a final decision maker, in our case that was the physician sponsor of the investigational device exemption. Similarly, during surgery, multiple team members independently validated the execution of the plan, but the study neurosurgeon was responsible for all final decision making. Given the critical nature of array placement in generating the expected sensation and the difficulty of a revision surgery, multiple redundant confirmations that the plan was being followed were critical to successful targeting of ICMS, particularly at this early stage of BCI development. Clinical translation will ultimately require a streamlined process, likely facilitated by improvements in imaging techniques and surgical approaches that improve surgical planning, for instance through limiting brain shift ^59^.

### Limitations

While the mediolateral somatotopy was identifiable through imaging, we were unable to identify the anteroposterior somatotopy, which dictates the distal aspect of the finger sensation. Prior intraoperative mapping studies indicated that proximal sensations occur during stimulation on the anterior bank of the post-central gyrus, while distal sensations occur during stimulation somewhere between the middle and posterior bank of the gyrus with substantial participant-to-participant variability^32^. Functional imaging with a 7T magnet revealed a similar pattern, but only in a subset of participants^60^. Distal sensations should presumably occur at the border between Brodmann’s areas 1 and 2 based on non-human primate work^20,23^, but the location of that border in the human brain is difficult to assess in vivo and appears to be highly variable from person to person^22,61^. In our five participants no clear relationship between the electrode location and how distal the evoked sensation was appeared. In other reports Herring et al. reported proximal sensations with anterior arrays^56^, while Fifer et al. report one array on the anterior bank of the post-central gyrus that evoked proximal sensations on its most anterior electrodes and more distal sensations on its posterior electrodes; however, another array positioned similarly evoked exclusively distal sensations^55^. Given this variability and the current difficulty with imaging proximal-distal within-digit somatotopy, the development of a reliable method for identifying the location of fingertip representation on the post-central gyrus would be beneficial to BCI-related surgical planning.

Another avenue for future work is to distribute the electrodes more broadly across somatosensory cortex to ensure more even coverage of sensations. Our implants consisted of two 2.4x4mm arrays of electrodes wired in a checkerboard pattern (Figure 3). Implanting additional, smaller arrays or otherwise distributing stimulation sites more uniformly through the finger representation in somatosensory cortex should improve the chances that all digits are represented evenly. The density of this distribution could scale up as the robotics, sensorization, and stimulation algorithms improve.

## Conclusion

We have developed and validated an array placement methodology that reliably led to usable evoked sensations of the fingers. While we implanted NeuroPort Microelectrode Arrays (Blackrock Neurotech, Salt Lake City, Utah), the imaging and planning methods are generalizable to new devices being developed for future clinical trials^62–65^. Across 5 participants, the expected somatotopic organization of little finger to thumb sensations progressing along the mediolateral axis was observed, though there is sufficient variability between participants to necessitate generating personalized functional maps. Variability is even more significant in the anteroposterior organization of the proximal and distal finger segments. At the current, early stages of BCI development, functional neuroimaging maps should be independently and carefully reviewed by multiple researchers and clinicians to ensure consensus prior to array implantation. With sophisticated functional maps and careful planning, researchers can be confident in their planned experiments and, eventually, clinicians will be able to reliably place clinical devices for BCI users.

## Disclosures

NH and RG served as consultants for Blackrock Neurotech, Inc, at the time of the study. RG is also on the scientific advisory board of Neurowired LLC. MB, JC, and RG received research funding from Blackrock Neurotech, Inc. though that funding did not support the work presented here. PW served as a consultant for Medtronic.

## Data Availability

All data produced in the present study are available upon reasonable request to the authors.

## Acknowledgments

We would like to thank the participants for all their extensive effort towards the success of this project. We would also like to thank Candida Ustine for assistance with MEG data collection. This work was supported by the National Institute Of Neurological Disorders And Stroke of the National Institutes of Health (NIH) under Award Numbers UH3 NS107714. Work at the University of Pittsburgh was developed with funding from the Defense Advanced Research Projects Agency’s (DARPA, Arlington) Revolutionizing Prosthetics program (contract number N66001-10-C-4056). The content is solely the responsibility of the authors and does not necessarily represent the official views of the NIH or DARPA.

## Online Methods

### Participants

Five participants enrolled in a multi-site clinical trial (registered on clinicaltrials.gov, NCT01894802 between 2014 and 2022 and provided informed consent prior to any experimental procedures. All procedures were approved by the Institutional Review Boards at the University of Pittsburgh or the University of Chicago. Table 2 shows the participant demographics. Participant C1 (male), in his 50s at the time of implant, presented with a C4-level ASIA D spinal cord injury (SCI) that occurred 35 years prior. Filament tests revealed spared deep sensation but diminished light touch in the right hand (detection thresholds ranged from 0.6 to 2.0 g across digit tips). Participant C2 (male), in his 60s at time of implant, presented with a C4-level ASIA D spinal cord injury (SCI), along with a right brachial plexopathy, that occurred 4 years prior. He was nearly insensate across the whole right hand (thresholds were 15-60g across digits 2-5 and >300g on D1). Participant P2 (male), in his 20s at the time of implant, presented with a C5 motor/C6 sensory ASIA B SCI that occurred 10 years prior to implant. He was insensate in the ulnar region of the hand (digits 3-5) on both the palmar and volar surfaces but retained both diminished light touch and deep sensation on the radial side (digits 1-2) (thresholds were 1.4 g to 8 g on the thumb and index, respectively, and 180 g on the middle digit). Participant P3 (male), in his 20s at the time of implant, presented with a C6 ASIA B SCI that occurred 12 years prior. He was insensate in the ulnar region of the hand on both the palmar and volar surfaces but retained diminished light touch and deep sensation on the radial side (thresholds were 0.07 g and 1.6 g on the thumb and index and 8 g on the middle digit). Participant P4 (male), in his 30s at the time of implant, presented with a C4-5 ASIA A SCI that occurred 11 years prior. He was insensate across his entire right hand (760 grams in all zones). As an inclusion criterion for the study, none of the participants retained functional motor control of the targeted hand.

### Scanning Procedures

Each participant underwent functional MRI (fMRI) and/or magnetoencephalography (MEG) scanning sessions. During scans, participants were asked to attempt to move individual fingers through flexion and extension on the right hand. Attempted movements (vs. motor imagery) were explicitly instructed to amplify activation^66^. Below, we will describe the task and analysis pipeline first for the fMRI scans and then MEG.

### fMRI Task Design

Participants were visually cued using E-Prime (v. 2.0 Psychology Software Tools, Pittsburgh, PA, USA) to attempt the movement of individual digits with their palm facing down towards the MRI table. Instructions were projected into the scanner bore. The movement cues were first-person perspective videos of an actor repeatedly performing flexion and extension movements of an individual digit at ∼0.5 Hz. Participants were instructed to attempt to follow along with the videos to the best of their ability. Only right-hand tasks were performed. All participants were right-handed prior to injury, and unable to physically execute the task post-injury.

Participants completed a travelling wave paradigm to map digit selectivity in S1, which has been shown to be effective for individuals with spinal cord injuries^45^. The paradigm involves moving digits in sequence. Each 9 second digit movement block was immediately followed by a movement block of a neighboring digit. We used two different sequence cycles: forward and backward. The forward sequence cycled through the digits from thumb to pinky. The backward sequence cycled from pinky to thumb. Prior to starting a block, participants were shown text instruction detailing whether the block would be a forward sequence block or a backward sequence block. Each run consisted of 8 repetitions of each sequence. In addition, there were 9 seconds of fixation (no movement) placed at the beginning and end of each run. A run lasted 6 minutes and 20 seconds. The total number of runs varies for each participant (5-8), with an equal number of forward and backward runs (besides P4 with 2 backward runs and 3 forward runs).

### fMRI Data Acquisition

The MRI scanner and sequence parameters varied across participants and are shown in Table 3.

**Table 3.** MRI scanner information across participants. FOV=field of view; TR=repetition time; TE= echo time. D1-D5 reflects digits 1 through 5: D1=thumb, D2=index, D3=middle, D4=ring, and D5=little.

|  | **C1** | **C2** | **P2** | **P3** | **P4** |
| --- | --- | --- | --- | --- | --- |
| **Scanner Location** | Chicago | Chicago | Pittsburgh | Pittsburgh | Pittsburgh |
| **Scanner** | 3T Philips Achieva | 3T Philips Achieva | 3T Siemens TrioTim | 3T Siemens Prisma | 3T Siemens Prisma |
| **Head Coil** | 32-channel | 32-channel | 32-channel | 64-channel | 64-channel |
| **T1 sequence parameters** | Voxel Size: 1mm isotropic<br>TR: 7.4s; TE: 3ms; TI: 1s | Voxel Size: 1mm isotropic<br>TR: 7.4s; TE: 3ms; TI: 1s | Voxel Size: 1mm isotropic;<br>TR: 2.3s; TE: 3ms; TI: 1s | Voxel Size: 1mm isotropic;<br>TR: 2.4s; TE: 3ms; TI: 1s | Voxel Size: 1mm isotropic;<br>TR: 2.4s; TE: 3ms; TI: 1s |
| **Functional sequence parameters** | Voxel Size: 2mm isotropic;<br>TR: 1.5s; TE: 30ms<br>252 volumes | Voxel Size: 2mm isotropic;<br>TR: 1.5s; TE: 30ms<br>252 volumes | N/A | Voxel Size: 2mm isotropic;<br>TR: 1.5s; TE: 30ms<br>252 volumes | Voxel Size: 2mm isotropic;<br>TR: 1.5s; TE: 30ms<br>252 volumes |
| **Number of functional runs** | 8 | 6 | N/A | 6 | 5 |
| **Digits with activity above z-threshold ($Z > 3.1$)** | D1-D5 | D1-D3 | N/A | D1-D4 | D1-D4 |

#### C1 & C2

MRI data were obtained using a 3T Philips Achieve dStream MRI scanner (Philips Medical Systems, Netherlands) and a 32-channel head coil at the University of Chicago MRI Research Center. A high-resolution T1-weighted structural MRI scan (3D-MPRAGE sequence: 1x1x1 mm voxel size, in-plane matrix size: 256 x 256 mm, 192 slices, TR = 7.4 s, TE = 3.1 ms, TI = 1 s, FA = 8°) was collected at the start of the session. Functional scans were acquired using a T2*-weighted EPI acquisition sequence (2 x 2 x 2 mm voxel size, in-plane matrix size: 96 x 96 mm, TR = 1.5 s, TE = 30 ms, FA = 74°). Sixteen slices with a slice thickness of 2 mm and no slice gap were oriented, using the participant’s 3D T1-view, such that it would be centered on the anatomical hand knob, prioritizing the dorsal portion of the brain. After the structural scan, 252 volumes were collected for each of the experiment runs (each lasting 6 min and 20 sec). In total the scanning session took approximately 60 min.

#### P2

MRI data were obtained using a 3T Siemens TrioTim MRI scanner (Siemens, Erlangen, Germany) and a 64-channel head coil at the University of Pittsburgh Medical Center. A high-resolution T1- weighted structural MRI scan (3D-MPRAGE sequence: 1x1x1 mm voxel size, in-plane matrix size: 256 x 240 mm, 160 slices, TR = 2.3 s, TE = 2.98 ms, TI = 1 s, FA = 90°) was collected. fMRI sequences were not collected for P2 due to incompatibility with implanted hardware and participant discomfort with the scanner environment.

#### P3 and P4

MRI data were obtained using a 3T Siemens Prisma MRI scanner (Siemens, Erlangen, Germany) and a 64-channel head coil at the University of Pittsburgh Medical Center. A high-resolution T1- weighted structural MRI scan (3D-MPRAGE sequence: 1x1x1 mm voxel size, in-plane matrix size: 256 x 256 mm, 192 slices, TR = 2.4 s, TE = 3.1 ms, FA = 8°) was collected at the start of the session. Functional scans were acquired using a T2*-weighted EPI acquisition sequence (2 x 2 x 2 mm voxel size, in-plane matrix size: 94 x 110 mm, TR = 1.5 s, TE = 30 ms, TI = 1 s, FA = 90°). Twenty-four slices with a slice thickness of 2 mm and no slice gap were oriented, using the participant’s 3D T1- view, such that it would be centered on the anatomical hand knob and prioritizing the dorsal portion of the brain. After the structural scan, 252 volumes were collected for each of the experimental runs (each 6 min 20 sec). In total the scanning session took approximately 60 min.

### fMRI Analysis

Functional MRI data processing was carried out using FMRIB’s Expert Analysis Tool (FEAT; Version 6.0), part of FSL (FMRIB’s Software Library, Oxford, UK), in combination with custom bash, Python (version 3) and Matlab scripts (R2019b, v9.7, The Mathworks Inc, Natick, MA, USA). Cortical surface reconstructions were produced using FreeSurfer (v. 7.1.1; and Connectome Workbench (humanconnectome.org) software.

### fMRI Preprocessing

The following pre-statistical processing was applied: motion correction using MCFLIRT^67^, non-brain removal using BET^68^, spatial smoothing using a Gaussian kernel of FWHM 3mm for the functional task data, grand-mean intensity normalization of the entire 4D dataset by a single multiplicative factor, and high-pass temporal filtering (Gaussian-weighted least-squares straight line fitting, with σ = 90 s). Time-series statistical analysis was carried out using FILM with local autocorrelation correction^69^. The time series model included trial onsets convolved with a double γ HRF function; six motion parameters were added as confound regressors. Indicator functions were added to model out single volumes identified to have excessive motion (>.9 mm). A separate regressor was used for each high motion volume (deviating more than .9mm from the mean position). The average number of outlier volumes for an individual scan varied across participants: (C1: 0; C2: 151; P3: 0.6; P4: 10). Finally, the functional data for each individual scan run within a session were then registered to the participant’s structural T1 using FLIRT^67,70^.

C2 experienced claustrophobia during the fMRI scan that led to substantial head motion (motion outlier volumes reflect over 50% of scan volumes). For C2 alone, we opted to analyze the functional data without motion scrubbing, due to the majority of the motion outliers occurring during any movement attempt. While we opted to include the visualization of this data in the supplementary materials, we primarily focused on their MEG data to guide array implantation.

### fMRI Analysis

We applied a general linear model (GLM) using FMRI Expert Analysis Tool (FEAT) to each functional run. To capture digit selectivity, the activity for each digit, in each functional run, was modelled as a contrast against the sum of the activity of all other digits of the same hand. Then, data for each digit, across the travelling wave runs, were averaged in a voxel-wise manner. A fixed effects model with a cluster-forming z-threshold of 3.1 and family-wise error corrected cluster significance threshold of *p* < 0.05 was used to differentiate between digits. Since the goal was to implant electrodes in the cortical surface, fMRI maps were projected to the surface using workbench command’s volume-to-surface-mapping function which included a ribbon constrained mapping method.

### Cortical Surface Reconstruction

Structural T1 images were used to reconstruct the pial and white-grey matter surfaces using Freesurfer^71^. Surface co-registration across hemispheres and participants was done using spherical alignment. Individual surfaces were nonlinearly fitted to a template cortical surface, first in terms of the sulcal depth map, and then in terms of the local curvature, resulting in an overlap of the fundus of the central sulcus across participants.

### MEG Task Design

For the MEG scans, participant P2 viewed videos of a finger being touched with a q-tip and imagined feeling the touch. P4 and C2 attempted to move individual fingers to match the movements seen on a first-person video (P4), or as demonstrated in real-time by a researcher (C2). The pace of movement was 0.5 Hz for Pittsburgh and 0.33 Hz for Chicago. Between 60 and 150 trials were collected for individual fingers. Due to comfort and time constraints, not all fingers were evaluated. Table 4 shows the trials used for each participant’s analysis.

**Table 4.** Details about MEG tasks and analysis. Blank entries represent tasks not performed.

|  | Rate (Hz) | # of trials/digit | # of Trials Used for Analysis |  |  |  |  | Time of Peak Activation (ms) |  |  |  |  |
| --- | --- | --- | --- | --- | --- | --- | --- | --- | --- | --- | --- | --- |
|  |  |  | D1 | D2 | D3 | D4 | D5 | D1 | D2 | D3 | D4 | D5 |
| **C2** | 0.33 Hz | 150 | 91 | 117 |  | 121 |  | 555 | 493 |  | 585 |  |
| **P2** | 0.5 Hz | 60 | 55 | 54 |  |  | 51 | 660 | 545 |  |  | 408 |
| **P4** | 0.5 Hz | 100 |  | 74 |  |  |  |  | 391 |  |  |  |

### MEG Data Acquisition

MEG data was collected on a 306-channel system at the Center for Advanced Brain Magnetic Source Imaging (CABMSI) at UPMC Presbyterian Hospital (Elekta for P2, MEGIN Triux for P4) or at the Medical College of Wisconsin (Elekta Neuromag for C2). Participants were transferred from their wheelchairs to the MEG compatible chair and wheeled into the system. Special attention was given to physically supporting the patient, including placing their hand on a tray in their line of sight. Data were collected at 1000 Hz using standard MEG recording procedures. Head position was tracked during the study and linked to digitized points on the head. MEG data were preprocessed by manually removing bad channels prior to performing temporal signal-space separation (tSSS) with a 4 second buffer^72^.

### MEG Analysis

MEG analysis was performed with Brainstorm software (http://neuroimage.usc.edu/brainstorm/)^73^ following published procedures^47^. Trials were manually inspected and removed for abnormal amplitude due to artifacts or excessive head movement. The data were further filtered 1 – 25 Hz and averaged by finger for each participant. The head position data were used to locate the MEG sensor positions on participants’ T1 MRIs. Anatomical models were created using FreeSurfer. Subsequent forward models were created using the overlapping spheres analytical model with 15,002 dipole locations. Current density reconstruction was used to provide a distributed map of activity on the cortical surface. Specifically, a weighted minimum norm estimate with unconstrained dipole orientations was used to allow for gyral activity. The one second of data before the observed movement onset was used to estimate baseline noise covariance.

A time point approximately 500 ms after cue onset (i.e., the start of the video) was chosen to create current density maps. Specifically, the time of the peak of the average current density power in the post-central gyrus region of interest (ROI) was chosen between 350-700 ms. This created cortical maps that represent the peak somatosensory activity during the beginning of the movement.

### Region of Interest

An anatomical sensorimotor (M1-S1) ROI was defined in the FreeSurfer average template space based on probabilistic cytoarchitectural maps^61^. This ROI was then projected onto the individual brains via the reconstructed individual anatomical surfaces. We focused the anatomical ROI on just M1 and S1 by selecting all surface nodes with the highest probability for Brodmann areas 1, 2, 3, and 4^74^. Further, we restricted the ROI to just the area roughly representing the hand, by excluding all surface nodes that had a distance greater than 2.5 cm from the anatomical hand knob^75^. We’ve provided a visualization of the digit selectivity maps masked with and without the ROI.

Finally, we used additional ROIs for visualization purposes in the “*Anatomical Hand Knob Proximity Analysis*”. For these ROIs, we loaded the FreeSurfer average cortical surface with the boundaries of either the BA1 or BA4 ROI, as defined by the Glasser atlas^76^. These ROIs were mapped onto individual anatomical surfaces.

### Activity Visualization

The fMRI and MEG activity were z-scored and masked with the M1-S1 ROI. Due to differences in modelling finger activity for each imaging paradigm (i.e., fMRI: digit selectivity; MEG: digit activity), different activity thresholds were applied. For the fMRI visualizations, we applied a minimum z-threshold of 3.1. Any digits that did not meet this statistical criterion were not included in the visualization. For the MEG visualizations, we applied a minimum z-threshold of 50% the maximum z-score across the cortex. The cortex over which the z-score was calculated did not include inferior and mesial areas since these areas do not contain sensorimotor activity. While activity below these thresholds may still be informative, thresholds were picked for standardization in the publication. The masked digit clusters were then mapped onto the participant’s cortical surfaces for visualization. To intuitively visualize all digits, we stacked the digit clusters such that the smallest cluster was the foreground overlay and the largest cluster was the underlay. The absence of digits in the MEG visualizations reflects the digit not being assessed (Table 4), while absence in an fMRI visualization reflects no supra-threshold activity for that digit (Table 3).

### Anatomical Hand Knob Proximity Analysis

A key question was whether implanting electrodes using the anatomical hand knob in the precentral gyrus as a guide would have been sufficient for identifying the location of digit- selective areas in S1. If true, an anatomically guided implantation would be much simpler. To calculate the distance along the post-central gyrus between digit activity and the anatomical hand knob, we implemented the following pipeline: 1) a neurosurgeon identified the precentral gyrus hand knob location on a T1w MRI scan; 2) we created a flattened version of the cortical surface for each participant; 3) the S1 homologue coordinate in the post-central gyrus was identified on the flat map; and finally 4) the distance from the S1 hand knob homologue coordinate to the peak functional activity for each digit was calculated (Supplementary Figure 1).

### Flattening the cortical surface

We generated flattened cortical surfaces using FreeSurfer commands. Our cutting strategy involved performing five cuts: one cut placed along the calcarine sulcus, three cuts equally spaced on the medial surface and one sagittal cut through the temporal pole (as described in https://freesurfer.net/fswiki/FreeSurferOccipitalFlattenedPatch). The flat maps were then created by using the *mris_flatten* function^61^. We then rotated the flat cortical surface of each participant, such that BA4 (defined using the Glasser atlas) was perpendicular to the x-axis. We then drew a horizontal line starting at the hand knob coordinate to the mid-point of BA1. This coordinate was used as the participant’s post-central gyrus hand knob homologue. This pipeline is visualized in Supplementary Figure 1.

### Computing distances to S1 hand knob homologue

To compute the distances between peak digit activity and the S1 hand knob homologue, we projected each participant’s thresholded digit activity onto the flat cortical surface. For fMRI, we only included digits with functional activity greater than a z-threshold of 3.1. With these thresholded digit maps, we then masked the activity to BA1. For each digit, we identified its peak activation coordinate (visualized in Supplementary Figure 1). Finally, we computed the distance along the mediolateral axis between each digit’s peak coordinate and the S1 hand knob homologue. In the results section, we report the minimum and maximum distance values across participants and digits.

### Implant Plan Generation

Functional sensory maps and motor maps (not discussed in this paper) were compiled into slides with both standard and flattened views of the left hemisphere, with a final slide showing only the standard view of the anatomical scan and two rectangles scaled to match the size of the arrays as inserted in the brain. These slides were distributed to seven or eight members of the research team, including neuroscientists, engineers, rehabilitative medicine clinicians, and neurosurgeons. Each person oriented and positioned the rectangles on the anatomical scan as they would like to see them placed in the surgery. All of the individual plans were compiled into a single set of slides and distributed back to the team members prior to a consensus meeting at which each person explained their reasoning. After full discussion of the individual plans, the team came to a final unanimous decision on a placement plan for the two arrays (Supplementary Figure 3), sometimes with an alternative location included in case one of the primary sites was discovered to be unusable during surgery.

### Surgical Navigation

Structural imaging and preoperative array locations were uploaded into StealthStation S8 (Medtronic, Minneapolis, MN), BrainLab (Curve, Munich, Germany), or ROSA (Zimmer Biomet, Warsaw, Poland) software. After induction of general anesthesia, the patient’s head was fixated with a Mayfield head clamp (Integra, Princeton, NJ) with an appropriate attachment for the given navigation system. Participant anatomy was registered to the presurgical imaging using face tracing, and registration accuracy was verified at multiple cranial landmarks. A surgical incision site was then planned using the neuronavigation software to ensure a craniotomy of sufficient size for the implantation of the array while also leaving space for the percutaneous pedestal connectors that were attached near the midline of the skull. Anatomic localization was again confirmed intraoperatively after the opening of the dura using the navigation system.

### ICMS

Stimulation surveys were used to determine where on the hand sensations were felt when each electrode was stimulated. Stimulation pulses consisted of a 200 µs cathodic phase followed by a 100 µs interphase and finally a half-amplitude 400 µs anodic phase. Pulses were delivered at 100 Hz for 1 second and varied in amplitude such that the percepts were reliable (usually at or above 60 µA). Electrodes were stimulated in a random order. The participants reported the location of the experienced sensation either verbally using a labeled hand map (Figure 5a) or by drawing the location on the map themselves using a touch screen interface. In addition to the sensation location, participants also reported what the sensation felt like. Participants could request repeated stimulation to help them discern the precise sensation. Surveys were repeated in each participant at least quarterly, but most electrodes showed little change over time^16^. On average, each electrode was tested 10, 4, 54, 16 and 3 times in C1, C2, P2, P3 and P4 respectively.

**Figure 5.**
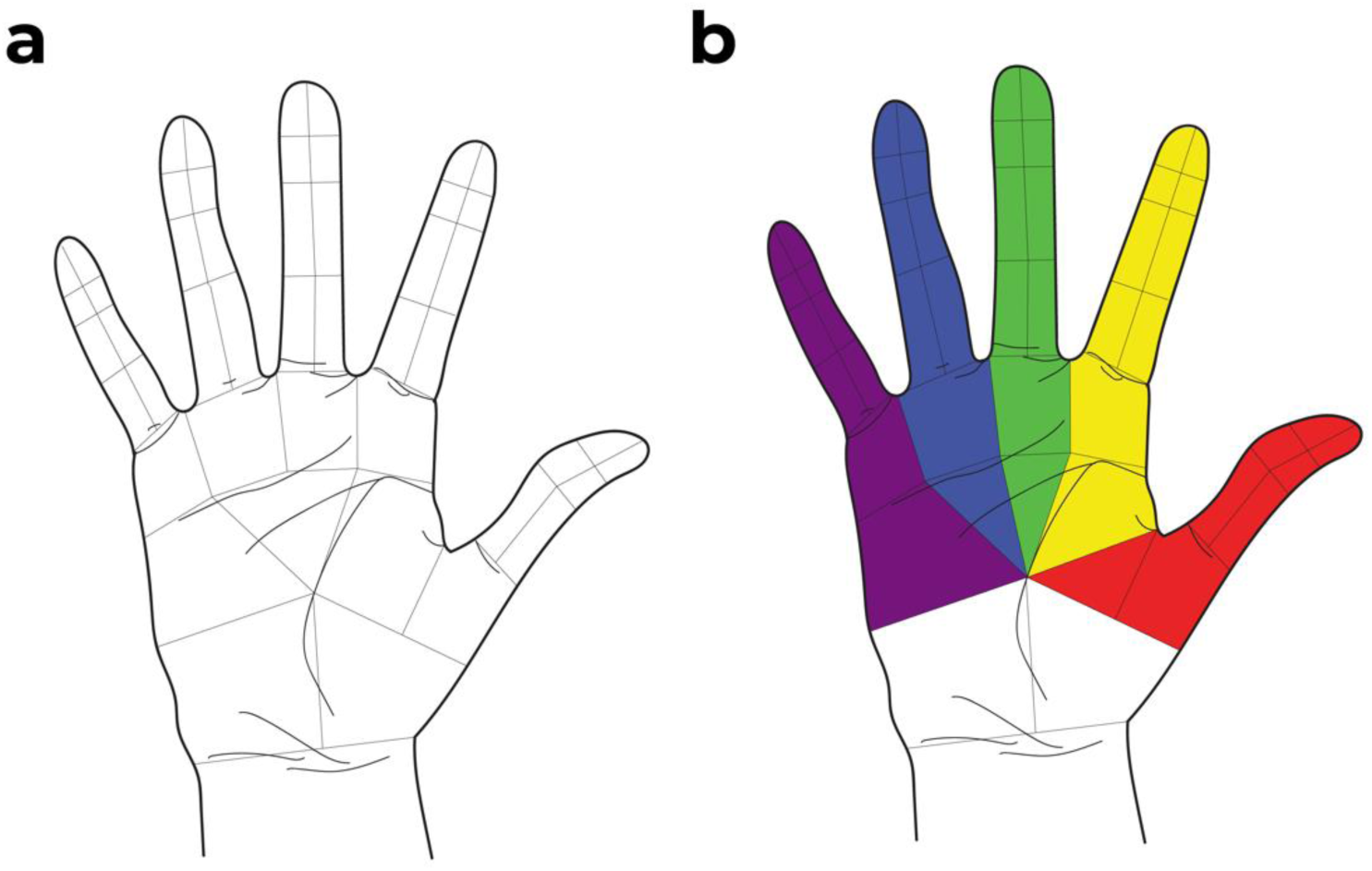
A hand map. a. Hand map that was used by the participants to report the experienced location of sensation in response to stimulation. b. The areas that were considered to correspond to each finger are indicated by individual colors.

To assess where each stimulation channel evoked a sensation, we first determined all pixels on the hand map that were enclosed by a drawn or verbally indicated hand area for each repetition of that channel. Each of these pixel groups was then assigned the label(s) of the hand map area(s) with which it overlapped (Figure 5b). Single electrodes often evoked sensations across multiple segments and digits. Hand segments that were reported in less than 30% of all times that a sensation was reported on that channel were deemed unreliable and excluded from further analysis. Furthermore, sensations reported on the lower side of the palm or wrist (P9-10 in Figure 5b) were excluded from further analysis.

### Comparing array-level sensations and neuroimaging functional maps

As a final analysis, we tested whether the neuroimaging digit activity under the implanted arrays accurately predicted the digit sensations reported when stimulating within the array. A crucial consideration for this analysis is the clear spatial differences in these estimates, namely the fMRI sequences had a voxel size of 2 mm^3^ and the arrays were 2.4 x 4 mm. For this reason, we opted to compare functional neuroimaging activity within any portion of the array boundaries to all evoked sensations from that array, as opposed to making comparisons on a channel-by-channel basis.

The positions of the stimulating arrays were determined using the surgical implant photos as a guide and 2.4x4 mm rectangles were visually registered on each participant’s cortical surface by a study team member. Using these rectangular areas as ROIs, we identified the digits with activity greater than the z-thresholds mentioned above. We considered these digits to be the predicted digits from the neuroimaging (see Figure 6 below).

**Figure 6.**
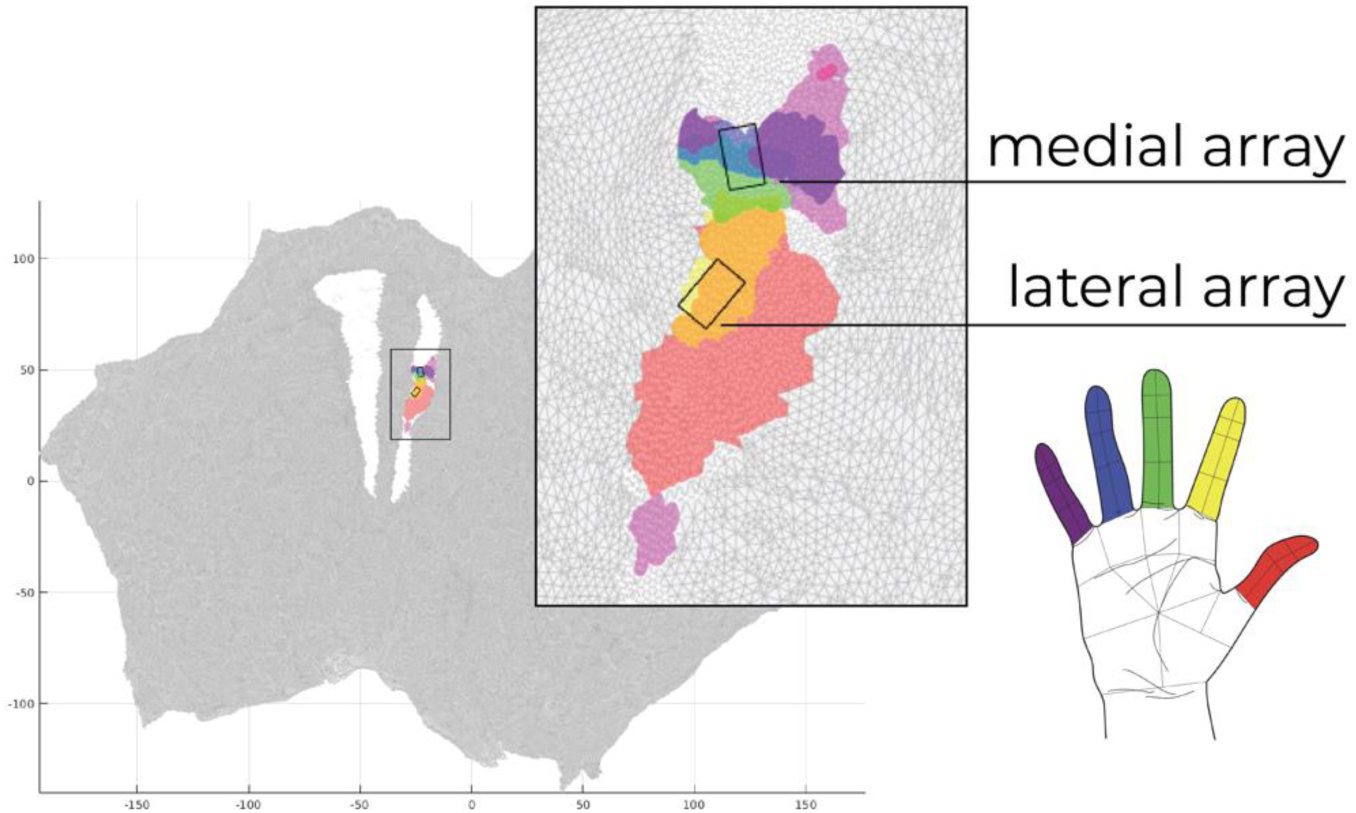
An example of functional neuroimaging activity within array locations for C1. The thresholded fMRI activity was projected on a flattened cortical map as shown. The estimated boundaries of Brodmann’s areas 4 and 1 are shown in white on the flattened map. The fMRI activity is colored based on the corresponding digit shown in the hand schematic. The locations of the stimulating arrays area outlined as black rectangles.

To calculate prediction accuracy, we compared the neuroimaging predictions to the digits where participants experienced a sensation evoked by stimulation of any electrode within an array. For each digit that was mapped under an array, we determined whether that array had at least one electrode that evoked a sensation in the mapped digit.

---

**Supplementary Figure 1.**
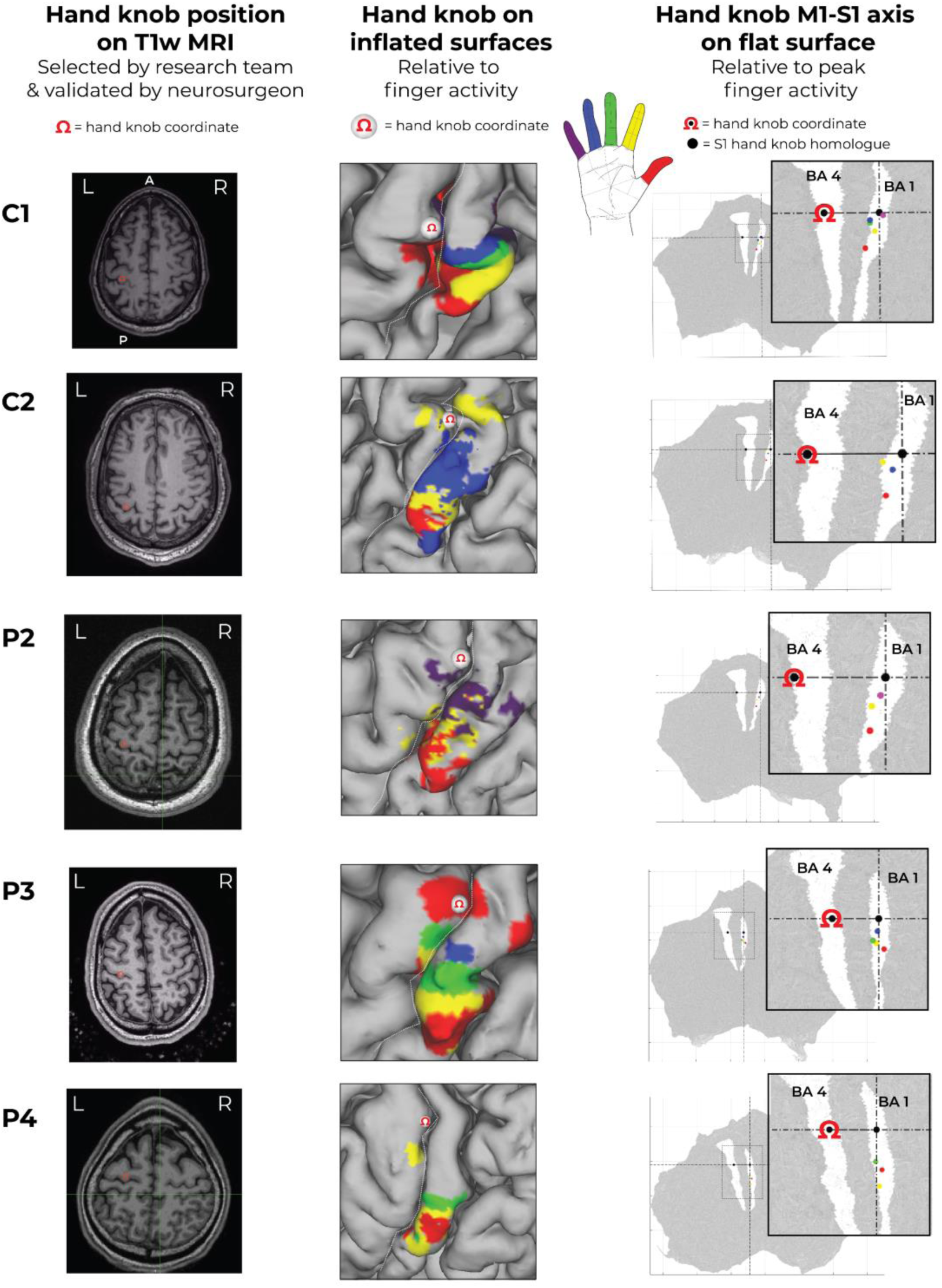
The locations of the peak functional activity on the post-central gyrus relative to anatomical hand knob. For each subject, a neurosurgeon selected the most likely hand knob (Ω) on the T1w scan. That location was transformed to the cortical surface model. Flattening the cortical model allowed for measurement of the lateral distance from the hand knob to the location of peak activity for each digit. In the right column, the estimated boundaries of Brodmann’s areas 4 and 1 are highlighted in white. Colored dots represent the location of the peak response for each digit.

**Supplementary Figure 2.**
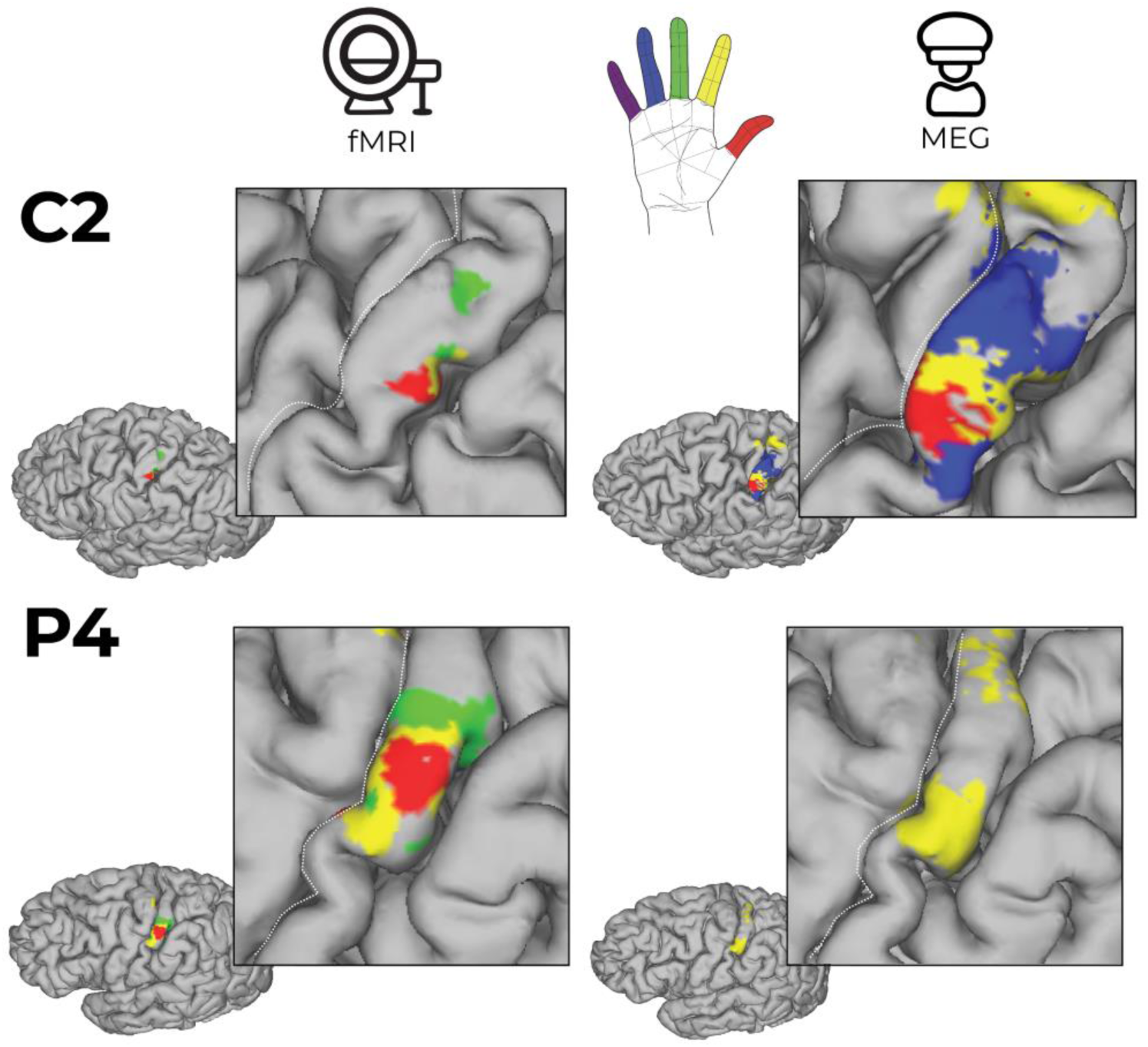
Comparison of fMRI and MEG results for two participants. Because of the low signal quality, C2’s fMRI data are thresholded at a lower z-score (2.3). Only index finger tasks were performed during the MEG for participant P4. All other annotations are the same as described in Figure 1.

**Supplementary Figure 3.**
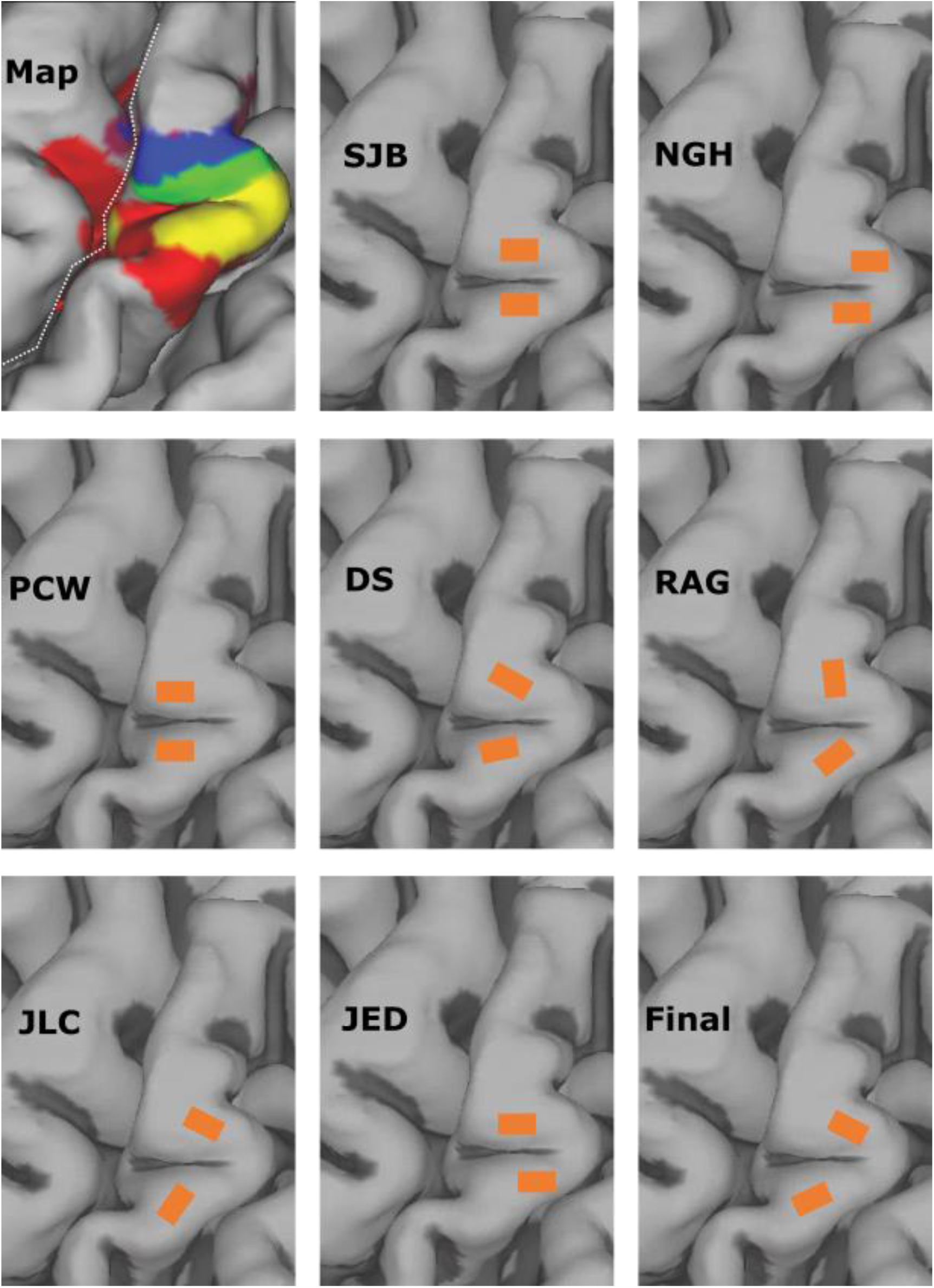
Development of consensus electrode array location for C1. The functional map shows activation for digits 1-5. Seven researchers generated independent plans for array placements based on function and structural imaging. The final placement plan was developed during a consensus meeting with the seven researchers plus the IDE sponsor-investigator present.

**Supplementary Table 1.** Representation of each digit by array. . Every array is shown and whether it overlaid a functional map of a given digit and what percentage of electrodes evoked a projected field on that digit. Percentages do not sum to 100, as projected fields for a single electrode could include multiple digits.

| Participant | Array | Digit | Map | Electrodes (%) | Participant | Array | Digit | Map | Electrodes (%) |
| --- | --- | --- | --- | --- | --- | --- | --- | --- | --- |
| C1 | Medial | D1 | No | 0.0 | P3 | Medial | D1 | **Yes** | **22.6** |
|  |  | D2 | No | 50.0 |  |  | D2 | **Yes** | **71.0** |
|  |  | D3 | **Yes** | **100.0** |  |  | D3 | **Yes** | **100.0** |
|  |  | D4 | **Yes** | **50.0** |  |  | D4 | No | 74.2 |
|  |  | D5 | **Yes** | **3.3** |  |  | D5 | No | 35.5 |
|  | Lateral | D1 | **Yes** | **28.6** |  | Lateral | D1 | **Yes** | **45.2** |
|  |  | D2 | **Yes** | **92.9** |  |  | D2 | **Yes** | **96.8** |
|  |  | D3 | No | 17.9 |  |  | D3 | No | 67.7 |
|  |  | D4 | No | 14.3 |  |  | D4 | No | 41.9 |
|  |  | D5 | No | 0.0 |  |  | D5 | No | 22.6 |
| C2 | Medial | D1 | **Yes** | **10.0** | P4 | Medial | D1 | No | 4.5 |
|  |  | D2 | **Yes** | **86.7** |  |  | D2 | **Yes** | **0.0** |
|  |  | D3 | -- | 93.3 |  |  | D3 | **Yes** | **0.0** |
|  |  | D4 | **Yes** | **86.7** |  |  | D4 | No | 13.6 |
|  |  | D5 | -- | 36.7 |  |  | D5 | No | 100.0 |
|  | Lateral | D1 | **Yes** | **15.4** |  | Lateral | D1 | **Yes** | **46.7** |
|  |  | D2 | **Yes** | **84.6** |  |  | D2 | **Yes** | **86.7** |
|  |  | D3 | -- | 96.2 |  |  | D3 | No | 100.0 |
|  |  | D4 | **Yes** | **92.3** |  |  | D4 | No | 73.3 |
|  |  | D5 | -- | 53.8 |  |  | D5 | No | 26.7 |
| P2 | Medial | D1 | No | 0.0 |  |  |  |  |  |
|  |  | D2 | **Yes** | **16.1** |  |  |  |  |  |
|  |  | D3 | -- | 61.3 |  |  |  |  |  |
|  |  | D4 | -- | 100.0 |  |  |  |  |  |
|  |  | D5 | **Yes** | **100.0** |  |  |  |  |  |
|  | Lateral | D1 | **Yes** | **70.0** |  |  |  |  |  |
|  |  | D2 | **Yes** | **93.3** |  |  |  |  |  |
|  |  | D3 | -- | 93.3 |  |  |  |  |  |
|  |  | D4 | -- | 86.7 |  |  |  |  |  |
|  |  | D5 | No | 43.3 |  |  |  |  |  |

## Notes

### Clinical Trial

NCT01894802

### Author Declarations

University of Chicago IRB, University of Pittsburgh IRB

